# Comparison of Ocular Surface and Tear Film Parameters in Smokers versus Non-Smokers in Palestine

**DOI:** 10.64898/2026.01.01.26343322

**Authors:** Mohammed Aljarousha, Abuimara Amer, Mohd Zulfaezal CHE Azemin

## Abstract

**Background:** Smoking is a significant global public health challenge, strongly associated with the rise of noncommunicable diseases and preventable deaths, particularly in conflict zones like Gaza.

**Aim:** To compare the different types of dry eye disease between smokers and non-smokers.

**Methods:** This cross-sectional study included 426 participants. Using a proportional stratified sampling approach, 80 participants were identified as smokers, which began in March 2022. An age- and gender-matched control group consisting of 80 participants was included for comparative analysis. Dry eye symptoms were assessed using the Arab-Ocular Surface Disease Index (Arab-OSDI) alongside several clinical tests. Dry eye was defined by an Arab-OSDI score of ≥ 13 and at least one positive clinical sign.

**Results:** Smokers had an average Arab-OSDI score of 17.09 ± 16.21 compared to 10.58 ± 10.65 in the control group (p = 0.046). The three ocular surface signs demonstrated statistically significant correlations in the expected directions (TBUT–STT: r ≈ 0.28; TBUT–CFS: r ≈ −0.24; STT–CFS: r ≈ −0.14; all p < 0.01). These inter-correlations were anticipated given the physiological relationships among the measures, and they were appropriately accounted for within the analytical model; therefore, they did not influence or confound the model estimates. Mild dry eye was observed in 13.6% of smokers and 7.4% of controls, while severe dry eye was present in 9.9% of smokers and 2.5% of controls. The overall frequency of dry eye was higher in smokers (25.9%) than in controls (11.1%) (p = 0.014). Aqueous deficient dry eye (ADDE) was more common among smokers (19.8%) compared to controls (8.6%; p = 0.041), as was mixed dry eye (p = 0.034).

**Conclusion:** Smokers exhibit a higher frequency of dry eye, with significantly elevated Arab-OSDI scores and higher incidences of ADDE and mixed dry eye associated with smoking.

## Background

Cigarettes contain over 4,000 harmful chemicals in both gas and particle forms that adversely impact the circulatory system, leading to vasospasm, platelet aggregation, and oxidative damage to lipids, proteins, and DNA within cells [1,2,3,4,5]. Smoking is a major risk factor for various cardiovascular, respiratory, and cancerous diseases [6]. It is also associated with several eye disorders, including primary open-angle glaucoma, optic neuritis, diabetic retinopathy, dry eye disease (DED), cataracts, Graves’ ophthalmopathy, age-related macular degeneration, and ocular inflammation [7,8]. Smoking reduces blood flow to the eyes and increases the risk of clot formation in ocular capillaries, depriving the eyes of vital nutrients needed for maintaining health [9]. Furthermore, passive exposure to tobacco smoke (commonly referred to as second-hand smoke) has been associated with ocular conditions such as hyperopia, dry eye disease, and cataracts [10,11].

Smoking increases the production of harmful free radicals while decreasing antioxidant levels in the blood, aqueous humour, and eye tissues, significantly raising the risk of oxidative damage to the eyes. Numerous eye-related conditions are linked to smoking, and this list continues to grow [12].

Hypothesis 1 confirmed in part: a longer (better) tear-film break-up time (TBUT) showed a robust negative association with symptom severity, while neither Schirmer’s test length (STT) nor corneal fluorescein staining (CFS) contributed uniquely when TBUT, smoking and age were controlled.

Hypothesis 2 not supported: age did not moderate any ocular-sign → OSDI paths; its influence was purely additive. The interaction terms (TBUT-Age, STT-Age, CFS-Age) were all non-significant.

The aim of this study is to compare the different types of dry eye disease between smokers and non-smokers.

## Methods

### Study design

This cross-sectional study included 426 participants. Using a proportional stratified sampling approach, 80 participants were identified as smokers, which began in March 2022. An age- and gender-matched control group consisting of 80 participants was included for comparative analysis. Participants were surveyed using a structured questionnaire to collect information about their history of and current exposure to passive smoking, as well as their usage of shisha and cigarettes. Shisha (also called waterpipe or hookah) is a device that heats flavoured or unflavoured tobacco; the smoke passes through water before inhalation. This matter was consistent with the methodology used in the Bukhari study [6]. Participants were recruited from the North Gaza Strip, Gaza City, the Middle Zone, and the South Gaza Strip, representing a total of five public and private eye-care facilities. These facilities included Al-Sahaba Medical Complex in the North Gaza Strip; Nasser Eye Hospital and the International Eye Hospital in Gaza City; Yafa Medical Hospital in the Middle Zone; and the European Gaza Hospital in the South Gaza Strip. The minimum required sample size was calculated as 384 using the formula n=(Z^2⋅p⋅(1-p)) /d^2, based on a 5% margin of error and a 5% type I error level. To enhance the reliability and precision of the findings, the final sample size was increased to 426 participants [13]. A total of 135 participants (31.7%) were selected from the South Gaza Strip, 62 (14.6%) from the Middle Zone, 147 (34.5%) from Gaza City, and 82 (19.3%) from the North Gaza Strip, proportionate to the population distribution in each region. The data collection occurred between March 15 and August 5, 2023.

### Ethical Approval

Ethical approval for this study was granted by the Palestinian Health Research Council Helsinki Committee (approval no. 883/21, dated 5 April 2021). All participants provided written informed consent before data collection began. For individuals unable to sign their own names, consent was confirmed through the signature of an independent witness.

### Inclusion and Exclusion Criteria

Inclusion Criteria:

1) Individuals aged 18 years and above. 2) Participants were matched by age within a ± 2-year range. 3) Resided in the study area during the data collection period (15 March–5 August 2023). 4) Capable of completing the demographic form, OSDI questionnaire, and clinical assessments. 5) Provided written informed consent.

Exclusion Criteria:

1) None of the participants in the control group were active smokers or had any history of passive smoke exposure—either at home or in the workplace. 2) participants with a history of wearing contact lenses, ocular conditions, using ocular medications, or having undergone ocular surgery or laser treatment within the past year. 3) Those with systemic diseases such as diabetes mellitus or hypertension as shown in **Figure 1**.

**Figure 1.**
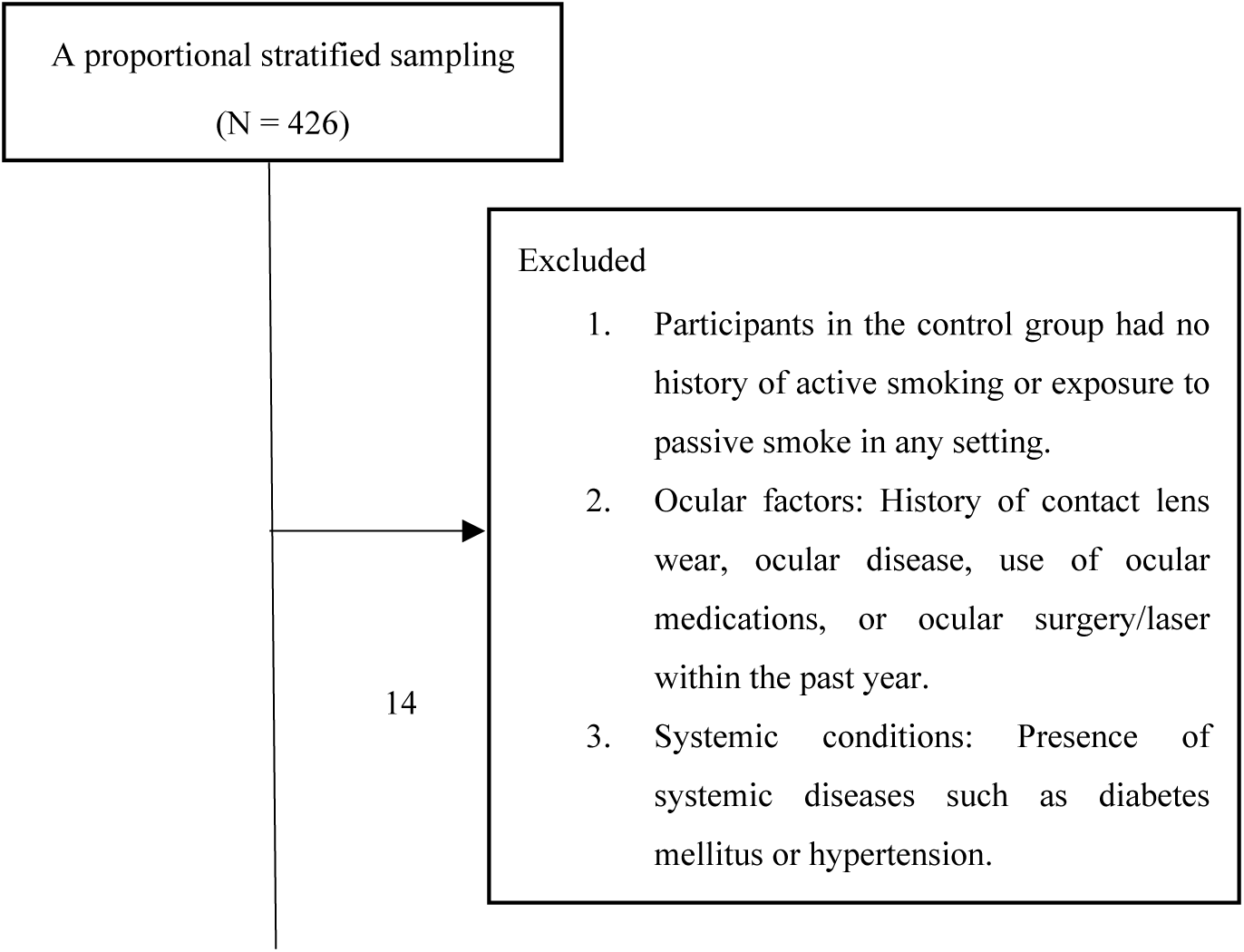
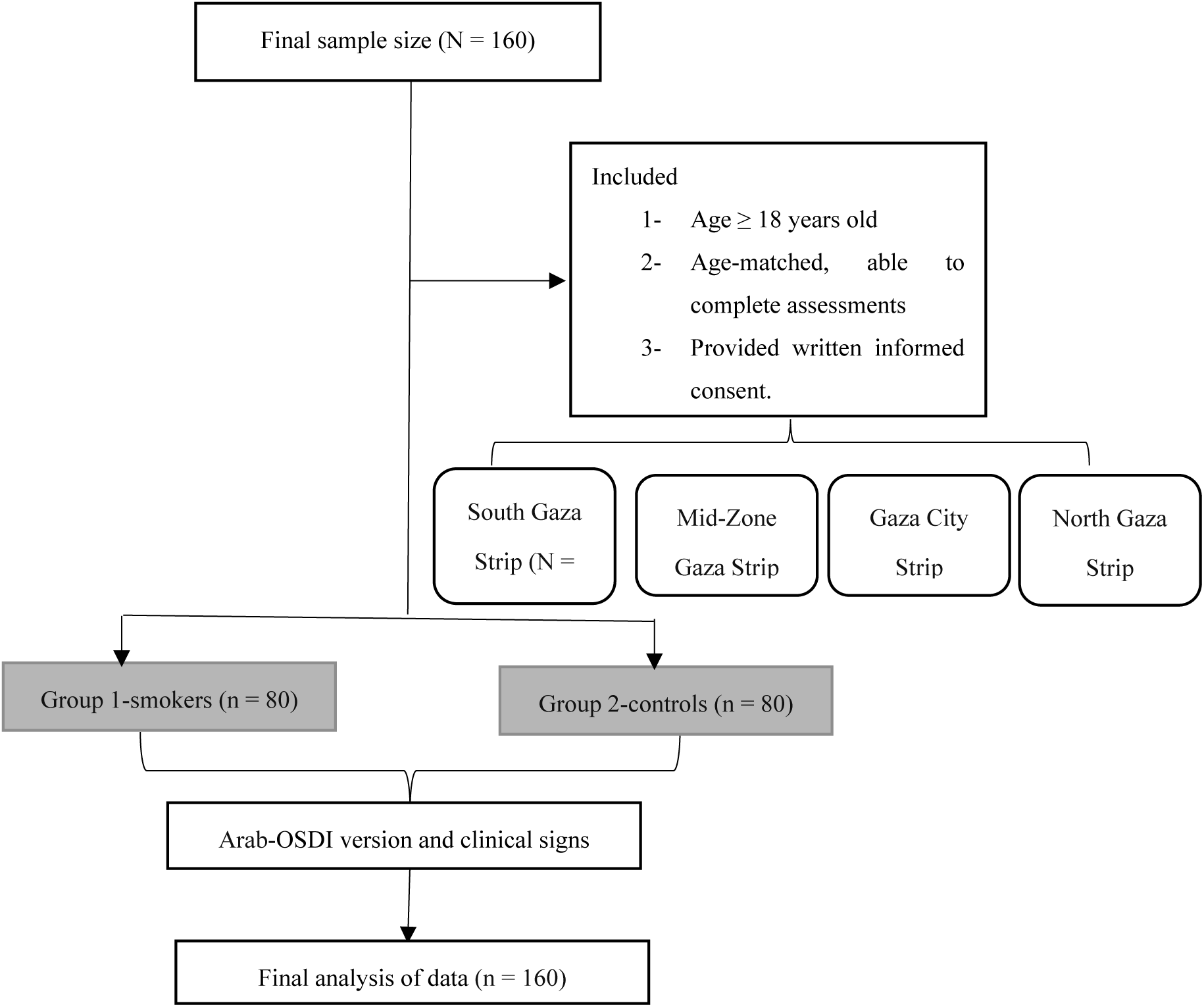
Flow diagram of the study.

### Outcomes and measures

The evaluation of dry eye symptoms in this study was conducted using the Ocular Surface Disease Index (OSDI) and several objective clinical tests. These tests included assessments of tear breakup time (TBUT), corneal fluorescein staining (CFS), Schirmer tear test with anesthesia (STT), tear meniscus height (TMH), meibomian gland dysfunction (MGD), Marx line staining (ML), and the lissamine green staining (LGS). The diagnostic criteria for dry eye disease was presented in **Table 1**.

**Table 1.** The criteria used to classify smokers and control groups were as follows:

| Diagnosis | Criteria |
| --- | --- |
| **DED** | Arab-OSDI score $\geq 13$<br><u>AND</u><br>Tear prism $< 0.2$ mm, MGDs $> 1$, ML $> 3.5$, LGS $\geq 1$, TBUT $< 5$ seconds, CFS $\geq 1$ or ST $< 15$ mm |
| **ADDE** | Arab-OSDI score $\geq 13$<br><u>AND</u><br>Tear prism $< 0.2$ mm or ST $< 15$ mm |
| **EDE** | Arab-OSDI score $\geq 13$<br><u>AND</u><br>MGDs $> 1$ or TBUT $< 5$ seconds |
| **Mixed DED** | Arab-OSDI score $\geq 13$<br><u>AND</u><br>Tear prism $< 0.2$mm or ST $< 15$mm <u>AND</u><br>MGDs $> 1$ or TBUT $< 5$ seconds |
| **DED symptom** | Arab-OSDI score $\geq 13$ |

DED: dry eye disease; ADDE: Aqueous deficient dry eye; EDE: Evaporative dry eye; OSDI: The ocular surface disease index; MGDs: Meibomian gland dysfunctions; ML: Marx line; LGS: conjunctival lissamine green staining; TBUT: Tear break up time test; CFS: corneal fluorescein staining; STT: Schirmer tear with anesthesia; $<$ : less than; ≥ : more than or equal.

### Techniques

#### Arab Ocular Surface Disease Index (OSDI)

The Arab-OSDI questionnaire consists of 12 questions that address ocular symptoms, vision-related issues, and environmental triggers [15]. Based on their scores, participants were categorised into normal (0–12), mild (13–22), moderate (23–32), and severe (33–100) groups. Those with an Arab-OSDI score of ≥13 were classified as having dry eye disease [14,15].

#### Invasive Tear Break-Up Time

TBUT was measured by placing a fluorescein sodium strip as a 0.25% solution moistened with a drop of 0.9% normal saline solution onto the bulbar conjunctiva of participants diagnosed with dry eye. Using a cobalt blue slit lamp beam, the time from the last blink to the appearance of random dark spots or streaks in the tear film was observed. Three readings were taken for each eye, and the average was used to determine the TBUT value in seconds. A participant with a TBUT of less than 5 seconds was diagnosed as having a positive clinical sign [16].

#### Corneal Fluorescein Sodium Staining

The fluorescein sodium dye (25% solution) was used to detect staining on the corneal epithelial surface, which appeared green under cobalt blue light. CFS was then assessed, with redness graded on a scale from 0 to 3: Grade 0 indicated no staining of the corneal epithelial surface; Grade 1 indicated mild staining confined to no more than one-third of the cornea; Grade 2 indicated moderate staining covering up to half of the cornea; and Grade 3 indicated severe staining covering more than half of the cornea [17].

#### Schirmer Tear Test

The STT was performed under local anesthesia by placing a strip of filter paper in the lower fornix laterally. After 5 minutes, the strip was removed, and the length of the wet portion was measured in millimeters. Participants with an STT measurement below 15 mm were diagnosed with insufficient aqueous deficient dry eye (ADDE) [18].

#### Tear Meniscus Height

TMH was measured by adjusting the slit lamp beam to a narrow setting and aligning it horizontally along the lower eyelid margin. Participants with TMH measurements below 0.2 mm were diagnosed with insufficient aqueous tear production [19].

#### Meibomian Gland Dysfunction

The assessment of meibomian gland (MG) obstruction involved a detailed examination of the eyelid margin using slit lamp biomicroscopy. The severity of MGD was classified on a scale from 0 to 4: Grade 0 indicated clear meibum; Grade 1 indicated colored meibum with normal consistency; Grade 2 indicated viscous meibum; Grade 3 indicated inspissated (thickened) meibum; and Grade 4 indicated a fully blocked meibomian gland [20].

#### Marx line staining

A lissamine green strip (1.5 mg per strip), moistened with unpreserved 0.9% saline, was applied to the lower eyelid to enhance visualization of the lid margin. The lower eyelid was divided into three zones—inner, middle, and outer—and each zone was graded on a 0–3 scale based on the secretion pattern: Grade 0 = no secretion reaching the gland orifices; Grade 1 = secretion touching the orifices; Grade 2 = secretion passing through all orifices; Grade 3 = secretion extending beyond the orifices onto the eyelid margin. The mean lid (ML) score was calculated, and values > 3.5 were considered abnormal [21] as shown in **Figure 2**.

**Figure 2.**
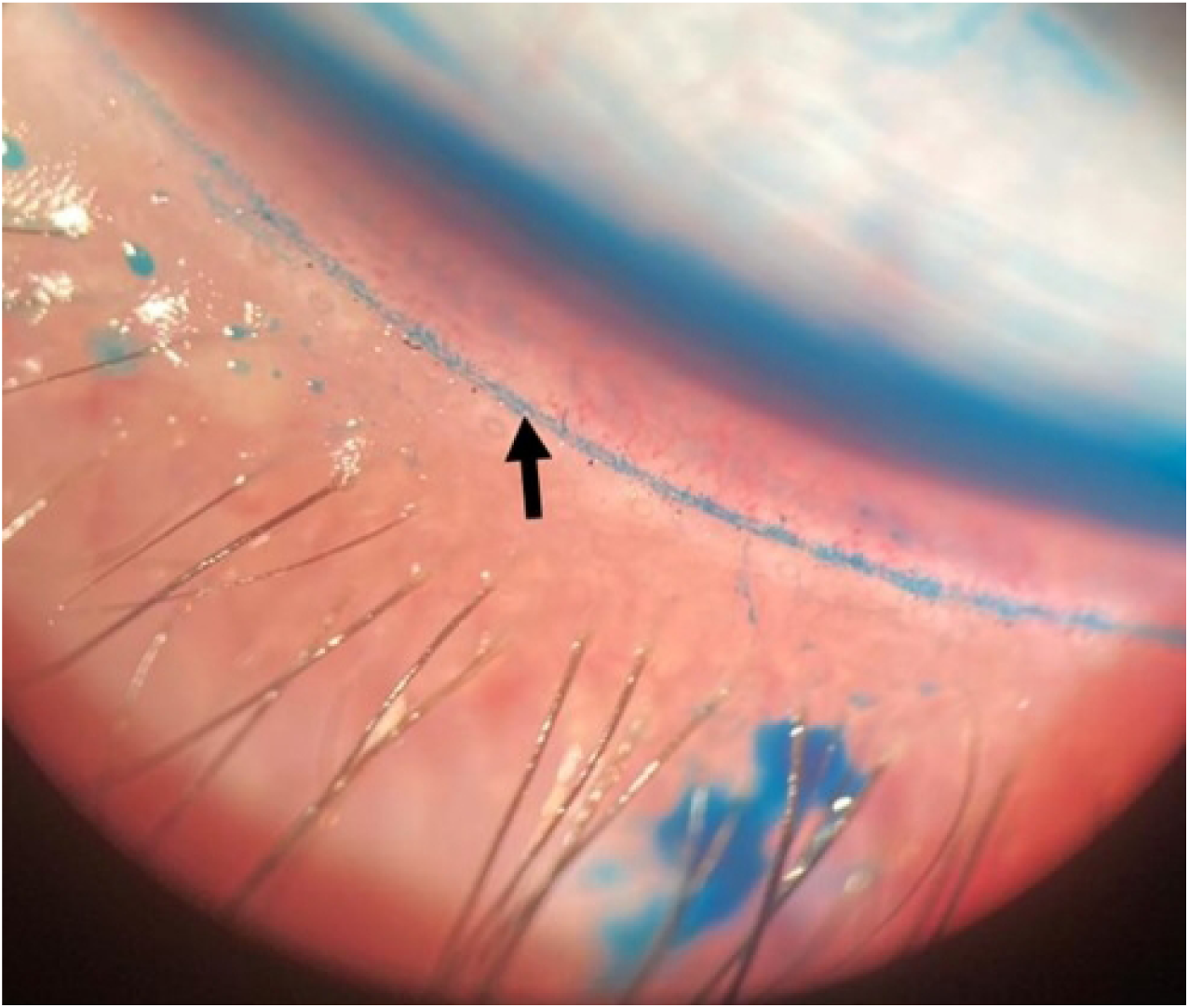
The lissamine green moistened the lower eyelid.

#### Lissamine Green Staining

The conjunctiva was assessed for lissamine green (1.5 mg concentration per strip) staining, with staining intensity graded on a scale from 0 to 5: Grade 0 indicated no staining; Grade 1 indicated minimal staining limited to 10 dots on the conjunctiva; Grade 2 indicated mild staining covering 32 dots; Grade 3 indicated moderate staining covering 100 dots; Grade 4 indicated marked staining covering 316 dots; and Grade 5 indicated severe staining covering more than 316 dots [22] as shown in **Figure 3**.

**Figure 3:**
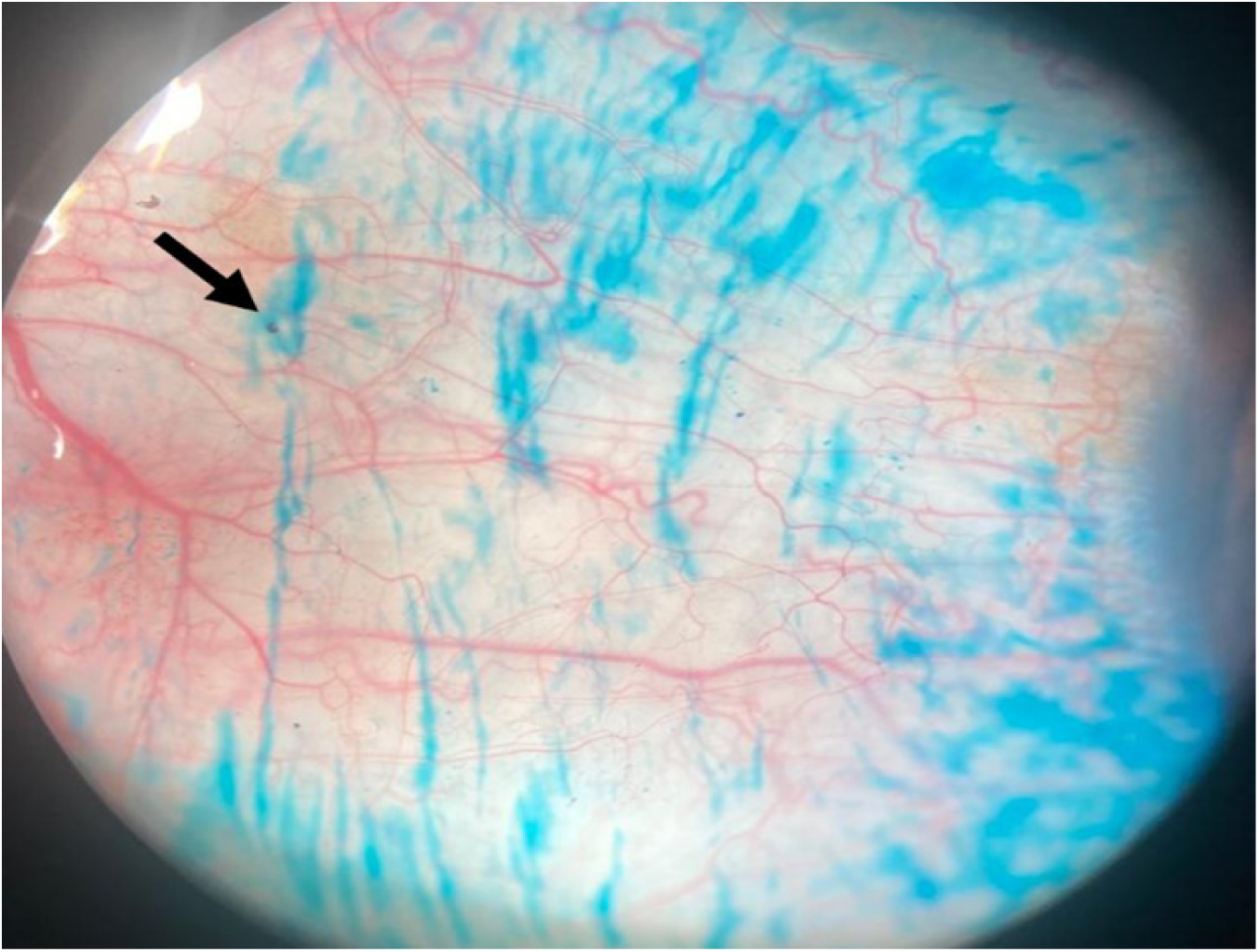
The conjunctiva lissamine green (1.5 mg concentration per strip)

#### Statistical analysis

Data were analyzed using IBM SPSS (version 26.0). The Shapiro-Wilk test was used to assess the normality of data distribution. The Chi-square test was employed to compare categorical variables, and the Wilcoxon signed-rank test was used to compare different types of dry eye between smokers and controls due to the non-normal distribution of the data. We fitted a moderated-mediation structural-equation model (SEM) in semopy 2.3.11.

## Results

### Baseline Characteristics of Smokers and Controls

In this study, 160 participants were enrolled (smokers n = 80; controls n = 80). The groups were comparable in age distribution—18–30, 31–40, 41–50, and > 50 years (smokers: 39 [48.8%], 16 [20.0%], 8 [10.0%], 17 [21.2%]; controls: 38 [47.5%], 18 [22.5%], 12 [15%], 12 [15%]; χ² = 0.448)— and in gender (smokers: 78 males [97.5%], 2 females [2.5%]; controls: 77 males [96.3%], 3 females [3.7%]; χ² = 0.665). No significant between-group differences were observed for region of residence (urban/rural; χ² = 0.261), type of residence (house/apartment/other; χ² = 0.595), working pattern (daytime fixed/rotating shifts/unemployed; χ² = 0.141), dietary pattern (vegetarian/meat/balanced; χ² = 0.181), use of heating and cooling systems (always/sometimes/not at all; χ² = 0.414), sleep duration (5–7 vs. 8–10 hours; χ² = 0.563), or use of dietary supplements or multivitamins (yes/no; χ² = 0.651) was presented in **Table 2**.

**Table 2.** Demographic data of smokers and control study participants.

| **Factors** | **Smokers group**<br>**n (%)** | **Control group**<br>**n (%)** | **Pearson Chi-square ($\chi^2$)** |
| --- | --- | --- | --- |
| **Age groups** |  |  | 0.448 |
| 18 - 30 (years) | 39 (48.8) | 38 (47.5) |  |
| 31 - 40 (years) | 16 (20.0) | 18 (22.5) |  |
| 41 - 50 (years) | 8 (10.0) | 12 (15.0) |  |
| > 50 (years) | 17 (21.2) | 12 (15.0) |  |
| **Gender** |  |  | 0.665 |
| Male | 78 (97.5) | 77 (96.3) |  |
| Female | 2 (2.5) | 3 (7.3) |  |
| **Region of residence** |  |  | 0.261 |
| Urban | 75 (93.8) | 71 (88.8) |  |
| Rural | 5 (6.3) | 9 (11.3) |  |
| **Residence** |  |  | 0.595 |
| House | 39 (48.8) | 39 (48.8) |  |
| Apartment | 41 (51.2) | 40 (50.0) |  |
| Others | 0 (0.0) | 1 (1.3) |  |
| **Working pattern** |  |  | 0.141 |
| Daytime fixed | 28 (35.0) | 74 (92.4) |  |
| Rotating shift work | 28 (35.0) | 5 (6.3) |  |
| Unemployed | 24 (30.0) | 1 (1.3) |  |
| **Menu** |  |  | 0.181 |
| Vegetarian | 20 (25.0%) | 16 (20.0%) |  |
| Meat | 18 (22.5%) | 8 (10.0%) |  |
| Balanced meals | 42 (52.5%) | 56 (70.0%) |  |
| **Heating and cooling system use** |  |  | 0.414 |
| Always | 6 (7.5%) | 11 (13.8%) |  |
| Sometimes | 42 (52.5%) | 8 (10.0%) |  |
| Not at all | 32 (40.0%) | 61 (76.3%) |  |
| **Sleeping hours** |  |  | 0.563 |
| 5 - 7 hours | 58 (72.5%) | 62 (78.0%) |  |
| 8 - 10 hours | 22 (27.5%) | 18 (22.0%) |  |
| **Dietary supplements (or multivitamins) use** |  |  | 0.651 |
| Yes | 18 (22.5%) | 23 (28.8%) |  |
| No | 62 (77.5%) | 57 (73.2%) |  |

### Associations and Interaction Impacts in the Structural Model

The three ocular signs inter-correlated in the expected directions (TBUT–STT r ≈ .28; TBUT–CFS r ≈ −.24; STT–CFS r ≈ −.14; all p < .01), but these covariances are accounted for in the model as shown in **Figure 4**. Longer TBUT was associated with lower OSDI, while smoking and older age were associated with higher OSDI. STT and CFS showed no unique effects once other variables were controlled. TBUT, STT and CFS did not predict smoking status; thus, indirect pathways via smoking were negligible. None of the interaction terms (TBUT×Age, STT×Age, CFS×Age) were significant; age acted additively rather than conditionally. In this sample, tear-film stability (TBUT) emerged as the key ocular sign linked to symptoms. Smoking and older age independently elevated symptom severity, but age did not alter the strength of the sign–symptom relationships, and smoking did not mediate them. These results prioritize TBUT and smoking status as clinically meaningful targets when interpreting or managing dry-eye symptoms.

**Figure 4.**
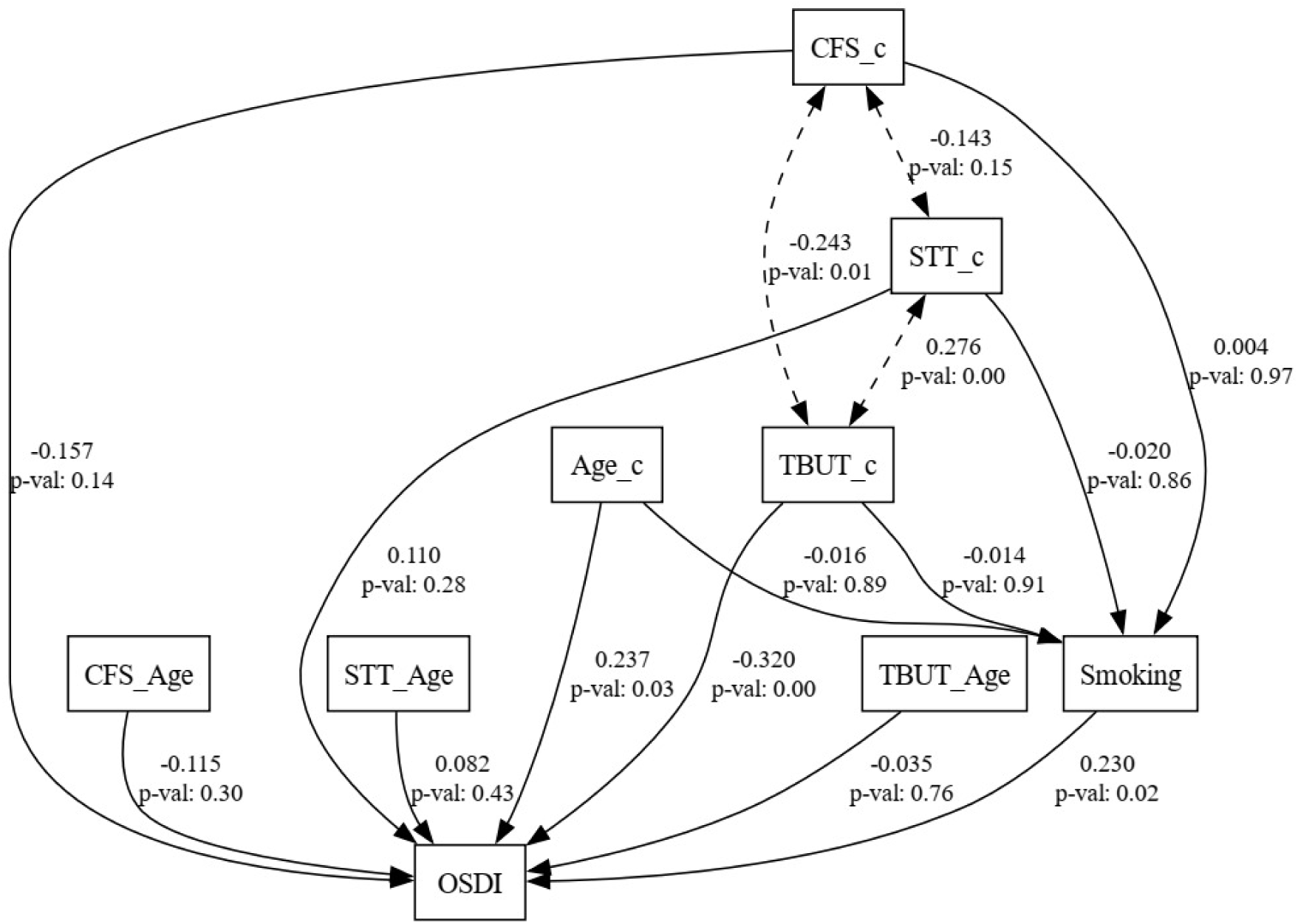
A path model was fitted in which tear-film break-up time (TBUT), Schirmer’s test length (STT) and corneal fluorescein staining (CFS) were entered simultaneously as predictors of dry-eye symptoms (OSDI), with smoking status specified as a mediator and age category as a moderator.

### Differences in OSDI Between Smokers and Controls Groups

As presented in **Table 3**, the Arab-OSDI scores, which measure dry eye symptoms, were higher in the smoking group compared to the control group, with mean ± SD values of 17.09 ± 16.21 for smokers and 10.58 ± 10.65 for controls (p = 0.046). In the smoking group, 21.0% had normal Arab-OSDI scores, compared to 35.8% of the control group. Mild Arab-OSDI scores were observed in 13.6% of smokers and 7.4% of controls, while moderate Arab-OSDI scores were similar between the two groups at 4.9%. Severe Arab-OSDI scores were more prevalent in smokers (9.9%) than in controls (2.5%), as depicted in Figure 2. A statistically significant difference in the severity of Arab-OSDI scores between smokers and controls was confirmed using the nonparametric Wilcoxon signed ranks test (p = 0.025).

**Table 3.**
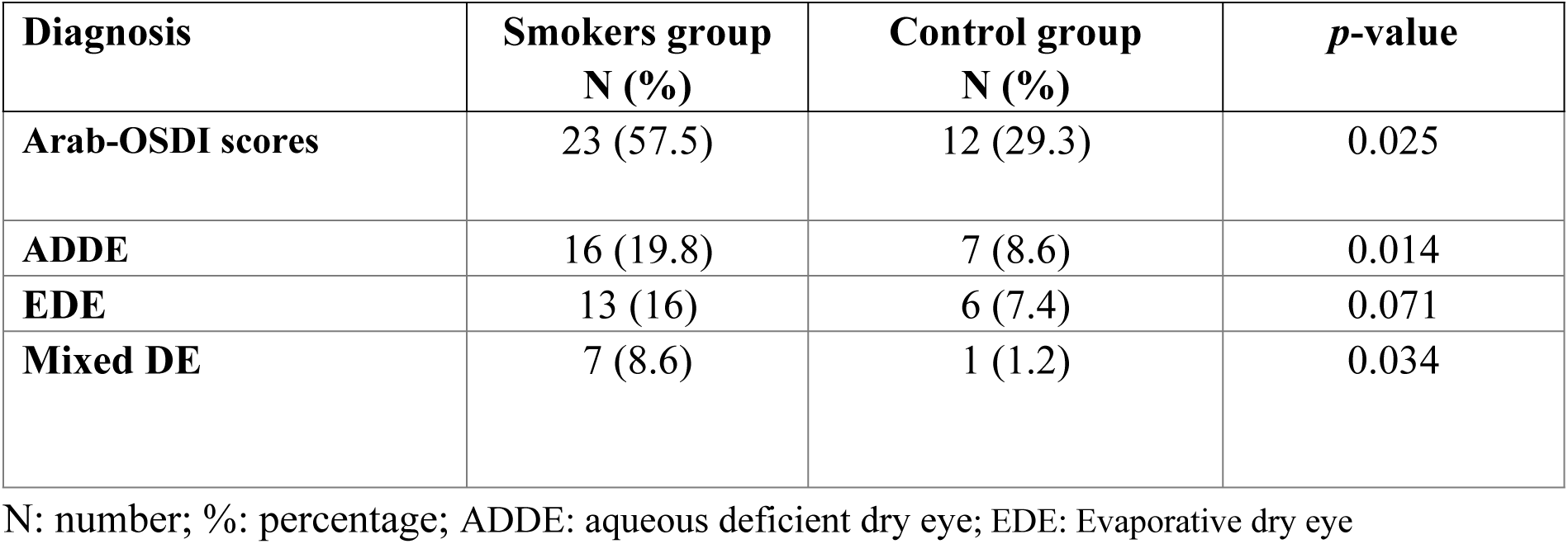
Comparison of different types of dry eye between the smoking group and control group.

| **Diagnosis** | **Smokers group**<br>**N (%)** | **Control group**<br>**N (%)** | ***p*-value** |
| --- | --- | --- | --- |
| **Arab-OSDI scores** | 23 (57.5) | 12 (29.3) | 0.025 |
| **ADDE** | 16 (19.8) | 7 (8.6) | 0.014 |
| **EDE** | 13 (16) | 6 (7.4) | 0.071 |
| **Mixed DE** | 7 (8.6) | 1 (1.2) | 0.034 |

N: number; %: percentage; ADDE: aqueous deficient dry eye; EDE: Evaporative dry eye

### Distribution of Dry Eye Types Among Smokers and Non-Smokers

The overall prevalence of dry eye was 57.5% in smokers and 29.3% in controls (p = 0.025). The occurrence of ADDE was significantly higher in the smoking group (19.8%) compared to the control group (8.6%), resulting in an 11.2% difference with a p-value of 0.041. Among smokers, the frequency of evaporative dry eye (EDE) was 16.0%, based on an Arab-OSDI score of ≥ 13 and meibomian gland dysfunction (MGD) score > 1 or tear breakup time (TBUT) < 5 seconds. Although this was 7.0% higher than in the control group, the difference was not statistically significant (p = 0.071), as indicated in **Table 4**. Additionally, the prevalence of mixed DE (both ADDE and EDE) was higher in smokers (8.6%) compared to controls (1.2%; p = 0.034).

## Discussion

Dry eye disease is a widespread condition characterized by unstable tear film, increased osmolarity, chronic inflammation, and nerve sensation abnormalities, ultimately leading to damage on the eye’s surface [23]. Smoking is recognised as a significant risk factor for dry eye. While smoking is widely known to contribute to various chronic diseases, its specific impact on dry eye remains less clear [24]. Previous studies, such as the Blue Mountains Eye Study, have reported a significant association between cigarette smoking and the development of dry eye [25]. Other research has similarly found that smoking increases the risk of dry eye [26–29]. Our study found a higher prevalence of dry eye among smokers compared to non-smokers, which is consistent with findings from Mohidin and Jaafar [23], who reported a significant link between tear film instability and smoking. Aziz et al. [3] noted that 34% of smokers experienced DE, and Lee et al. [17] found a connection between current smoking and dry eye symptoms. This may be due to smoking reducing the sensitivity of nerves on the ocular surface, decreasing awareness of symptoms.

The severity of dry eye symptoms was assessed using the Arab-OSDI score, with our findings aligning with those of Bhutia et al. [4], who also found a higher proportion of severe OSDI scores among smokers. While many studies have noted a correlation between smoking and dry eye disease, our study is the first to document the prevalence of different types of dry eye in both smoking and control groups in a Gaza sample. Smokers in our study had a higher prevalence of ADDE compared to controls, with significant differences in Arab-OSDI scores, TMH, and STT between the two groups. Symptoms of dry eye accompanied by decreased TMH or STT values indicate reduced basal tear secretion by the accessory lacrimal glands. Some research [31,32,33] suggests that smoking can damage the eye’s surface, leading to a decrease in goblet cell density and squamous metaplasia. However, other studies, such as those by Thomas et al. [34] and Tsubota et al. [35], found no significant differences in Schirmer’s test or TMH between smokers and non-smokers, likely due to variations in smoking habits and environmental factors.

In our study, smokers exhibited a higher prevalence of dry eye symptoms with shorter TBUT values or MGD compared to non-smokers. However, no significant difference in EDE was observed between the two groups [36]. A study in China similarly found a higher occurrence of MGD among smokers [37]. The detrimental effects of cigarette smoke on the tear film’s lipid layer likely contribute to the deterioration in TBUT. Other studies suggest that smoking increases the production of proinflammatory cytokines and reduces anti-inflammatory cytokines, leading to inflammation in the meibomian gland and contributing to MGDs, a primary cause of dry eye [38].

The current study revealed a significant difference in mixed dry eye, encompassing both ADDE and EDE, between smokers and non-smokers. To our knowledge, no previous study has documented the association between smoking and mixed dry eye or ML staining. Previous research has found that corneal staining, indicative of corneal epithelial damage, is significantly higher among smokers compared to non-smokers.

## Limitations

Our study did not include data on the classification of smokers based on their daily cigarette consumption, which is a limitation. Additionally, non-invasive tests such as TBUT or tear film osmolarity were not conducted. Finally, we did not investigate the association between dry eye parameters and the level of stress.

## Conclusion

This case-control study, matched for age and gender, found a significantly higher frequency of dry eye among smokers compared to controls. Smokers also had significantly higher mean Arab-OSDI scores. The most common type of dry eye among smokers was ADDE, followed by EDE and mixed dry eye disease. Further blinded investigations are recommended to better understand the relationship between smoking and dry eye disease.

## Declaration

### Author contributions

Mohd Zulfaezal Che Azemin and Sultan Alotaibi: Data analysis, Writing – result section. Mustafa Abdu and Mohd Zulfaezal Che Azemin: Introduction section and hypothesis analysis. Mohammed Aljarousha and Waleed Alghamdi: Data analysis, Methodology, Writing - review. Waleed Alghamdi and Mohammed Aljarousha: Writing – Discussion section. All authors contributed to the article and approved the submitted version. All authors have read and approved the final manuscript.

### Data Availability

The data that support the findings of this study are available from the corresponding author upon reasonable request.

### Ethics approval statement

Ethical guidelines for conducting research in conflict zones were strictly observed, including measures to protect participant confidentiality. Written informed consent was obtained from all participants.

### Clinical trial number

Not applicable.

## Data Availability

No

## Acknowledgements

The author(s) would like to thank Management and Science University for the technical support provided to publish the present manuscript.

